# Incidence and Risk Factors of Postoperative Delirium in Children with Congenital Heart Disease: A Retrospective Nationwide Inpatient Sample Database Study

**DOI:** 10.1101/2025.08.01.25332649

**Authors:** Ru-Gang An, Jin-Qing Tang, Jing Xu, Hao Xie, Bo-Fei Dong, Yan Wang, Zheng-Quan Tan, Ming-Qiang Zhou

**Author notes:** Corresponding author: Ming-Qiang Zhou, M.D, Department of Anesthesiology, The First People’s Hospital of Zunyi (The Third Affiliated Hospital of Zunyi Medical University), No.98, Fenghuang Road, Zunyi, Guizhou Province, 563000, China. These authors are correspondence authors.

## Abstract

**Background:** Postoperative delirium (POD) is an important neurocognitive complication following congenital heart disease (CHD) surgery in pediatric patients. While recognized for its association with adverse clinical outcomes, comprehensive population-based studies examining POD incidence and risk factors in this vulnerable population remain scarce.

**Methods:** Using the Nationwide Inpatient Sample (NIS) database (2010-2019), a retrospective study was performed of pediatric patients (<18 years) undergoing CHD-related surgeries. We evaluated demographics (race, sex, age), hospital characteristics (region of the hospital, type of admission, bed size of hospital, teaching status of hospital, type of insurance, location of hospital), mortality, length of stay (LOS), total charges, perioperative complications, and comorbidities.

**Results:** Among 37,535 CHD surgery patients identified, POD was diagnosed in 717 (1.9%). Children with POD exhibited higher comorbidity burdens, increased total medical costs, prolonged LOS and increased rates of in-hospital mortality (P < 0.0001). Independent predictors of POD included coagulopathy, drug abuse, fluid and electrolyte disorders, other neurological disorders, and paralysis. Additionally, POD was linked to in-hospital complications, including prolonged mechanical ventilation, acute renal failure, acute myocardial infarction, pneumonia, and respiratory failure.

**Conclusions:** Although the incidence of POD following CHD surgery in children is relatively low, investigating its predisposing factors is clinically valuable for optimizing patient management and improving outcomes.

## Introduction

Congenital heart disease (CHD) represents the most prevalent birth defect globally, affecting approximately 1% of live births (1, 2). Although advances in medical technology and surgical techniques have dramatically improved survival rates for children with CHD, making cardiac surgery a critical life-saving intervention, postoperative complications remain a significant concern(3, 4). Currently, approximately 40,000 pediatric cardiac surgeries are performed annually in the United States alone, with this number projected to increase due to enhanced diagnostic capabilities and improved healthcare accessibility (5).

Postoperative delirium (POD) is one of the most common and serious complications following CHD surgery in children(6, 7). POD is an acute neuropsychiatric syndrome characterized by alterations in attention, awareness, and cognition, typically emerging within hours to days after surgery (8). The reported incidence varies substantially, ranging from 10% to 80%, depending on diagnostic criteria and patient populations (9, 10). Evidence demonstrates that pediatric CHD patients developing POD experience significantly worse clinical outcomes, including longer periods of mechanical ventilation, extended durations of hospital and ICU admissions, and heightened healthcare expenses, and long-term cognitive impairment(11–13).Beyond financial burdens, POD has been shown to adversely affect patients, families, and healthcare systems, leading to higher mortality rates, progressive functional impairment, and reduced quality of life(14, 15).

The etiology of POD in pediatric CHD patients is multifactorial. Numerous risk factors have been recognized, such as the patient’s age, the severity of the CHD, and any cognitive impairments present before surgery(16–18). Additionally, factors such as cyanotic heart disease, developmental delays, and postoperative electrolyte disturbances have been linked to an increased risk of delirium(10, 19). Although research on POD in pediatric patients undergoing CHD surgery is increasing, many current studies are limited by small sample sizes, single-center designs, or inconsistent diagnostic criteria.

To address this gap, we leveraged the National Inpatient Sample (NIS) database to analyze over 30,000 pediatric CHD surgery cases surgeries. This study aims to elucidate the epidemiology of POD, identify key risk factors, and inform targeted prevention strategies to improve clinical outcomes in this vulnerable population.

## Materials and Methods

### Data source

This study employed data from the NIS, a principal database within the Healthcare Cost and Utilization Project (HCUP) supported by the Agency for Healthcare Research and Quality (AHRQ). As the largest all-payer database of hospital admissions in the United States, the NIS provides a comprehensive and stratified sample of over 1,000 hospitals, representing approximately 20% of annual hospitalizations nationwide. This database includes detailed information on patient demographics, hospital characteristics, length of stay (LOS), total hospitalization charges, payer types, in-hospital mortality, and diagnostic and procedural codes based on the International Classification of Diseases, Ninth and tenth revisions, Clinical Modification (IICD-9-CM and ICD-10-CM). For this retrospective analysis, de-identified data were extracted, ensuring compliance with ethical standards for the use of publicly available datasets. The NIS database’s robust sampling methodology and extensive coverage make it a reliable source for studying the incidence and risk factors of POD in children with CHD.

### Data access and ethics statement

This cross-sectional nationwide study analyzed fully de-identified patient data from the 2010-2019 US National Inpatient Sample (NIS) database (HCUP). Research data access occurred between 01/09/2024 and 15/12/2024 with no accessible protected health information at any phase. All NIS records were pre-anonymized prior to release with no identifiable personal information retained. The Institutional Review Board of The First People’s Hospital of Zunyi (The Third Affiliated Hospital of Zunyi Medical University) granted exemption from full ethical review and waived informed consent requirements, as researchers accessed only de-identified data in compliance with HCUP Data Use Agreement protocols.

### Data collection

The study analyzed 2010-2019 NIS data, and the ICD-9-CM and ICD-10-CM was utilized to identify patients who had surgery for CHD and to assess the incidence of POD. Study inclusion criteria encompassed patients under 18 years of age who underwent surgical intervention for CHD. POD was diagnosed using particular ICD-9-CM and ICD-10-CM diagnostic codes (Supplemental Table S1). Initially, 38,398 cases were identified. After excluding patients with missing data for key variables, such as gender, total hospital charges, length of stay, expected payment method, elective admission status, hospital bed size and death, the final analysis comprised 37,535 cases. (Fig 1)

**Fig 1.**
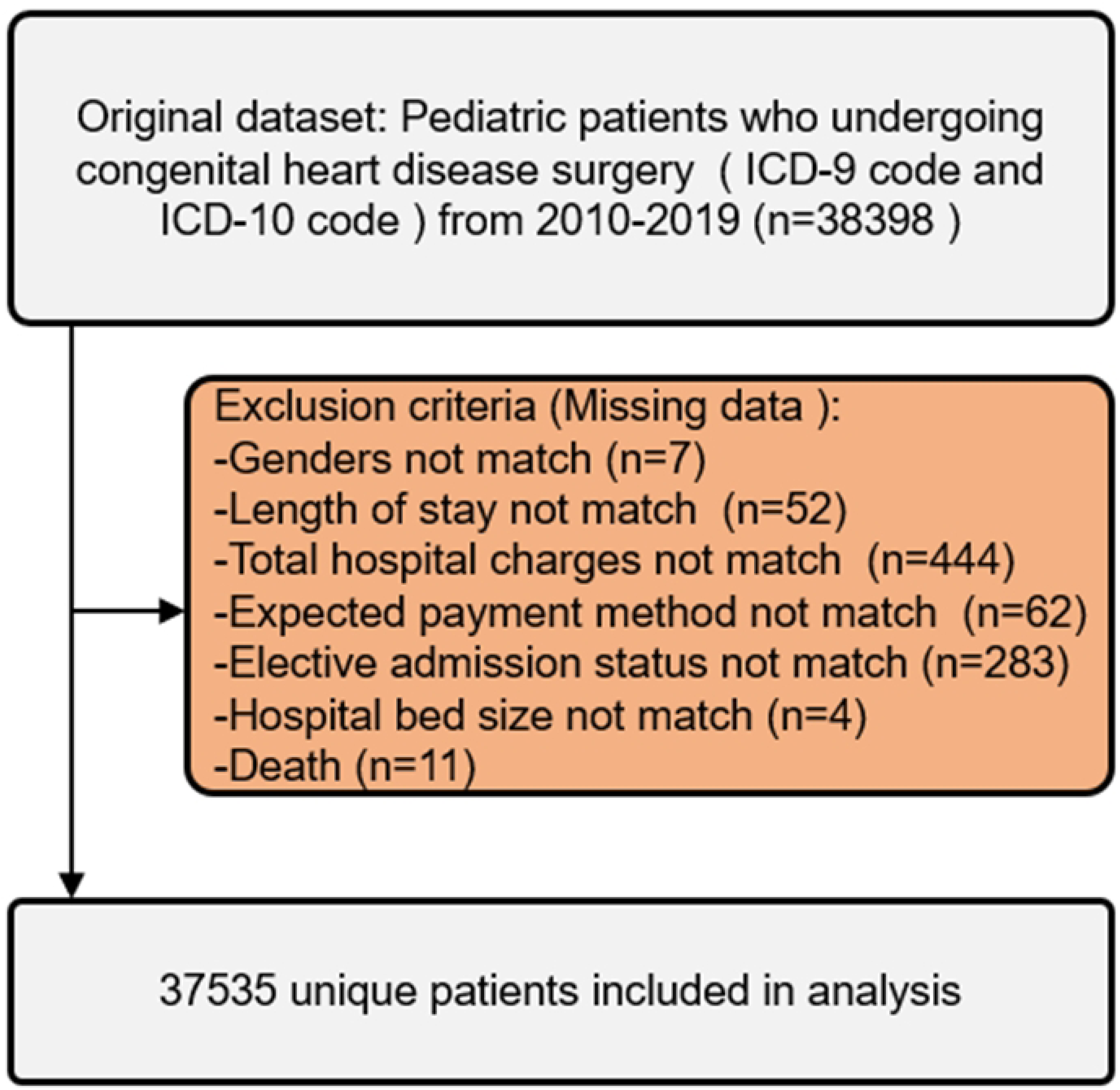
Exclusion process for pediatric patients undergoing congenital heart disease with postoperative delirium.

Patient demographics (including race, sex, and age), hospital characteristics (region of the hospital, type of admission, bed size of hospital, teaching status of hospital, location of hospital, type of insurance), and outcome measures such as length of stay (LOS), and total hospital charges (TOTCHG) and in-hospital mortality were extracted from the database. Additionally, information on preoperative comorbidities and perioperative complications was collected using ICD-9-CM and ICD-10-CM diagnostic codes (Table1). Perioperative complications included: pneumonia, blood transfusion, continuous invasive mechanical ventilation, urinary tract infection, acute renal failure, wound dehiscence/non-healing, acute myocardial infarction, arrhythmia, postoperative shock, and respiratory failure.

**Table 1.** Variables used in binary logistic regression analysis.

| Variables Categories | Specific Variables |
| --- | --- |
| Patient demographics | Age (0-2years/3-5years/6-12years/13-17years/), sex (male and female), race (White, Black, Hispanic, Asian or Pacific Islander, Native American and Other) |
| Hospital characteristics | Type of admission (non-elective, elective), bed size of hospital (small, medium, large), teaching status of hospital (nonteaching, teaching), location of hospital (rural, urban), type of insurance (Medicare and Medicaid, private insurance, self-pay, no charge, |
|  | other), region of the hospital (northeast, Midwest or north central, south, west) |
| Comorbidities | Deficiency anemia, chronic blood loss anemia, congestive heart failure, chronic pulmonary disease, coagulopathy, depression, diabetes (uncomplicated), diabetes (with chronic complications), drug abuse, hypertension, hypothyroidism, liver disease, lymphoma, fluid and electrolyte disorders, metastatic cancer, neurological disorders, obesity, paralysis, peripheral vascular disorders, psychoses, pulmonary circulation disorders, renal failure, valvular disease and weight loss |

### Data analysis

Statistical analysis was performed using IBM SPSS Statistics 29.0. Descriptive statistics were used to summarize patient demographics, hospital characteristics, and clinical outcomes. Continuous variables, such as age and length of stay (LOS), were analyzed using independent t-tests, while categorical variables, such as gender, race, and insurance type, were compared using chi-square tests. Univariate analyses evaluated potential associations between postoperative delirium POD and demographic/clinical variables. Independent predictors were discerned by means of stepwise binary logistic regression, incorporating patient demographics, hospital characteristics, and perioperative complications. Results are presented as adjusted odds ratios (OR) with corresponding 95% confidence intervals (CI). Given the substantial cohort size, a conservative significance threshold (α = 0.001) was adopted to minimize Type I error risk. All tests were two-tailed, and missing data were addressed through complete case analysis to maintain methodological rigor.

## Results

### The incidence of postoperative delirium in patients with CHD surgery

From 2010 to 2019, a total of 37,535 children undergoing congenital heart disease surgery were identified in the NIS database. Among these, 717 cases (1.9%) were diagnosed with postoperative delirium during hospitalization. The annual incidence of postoperative delirium exhibited slight fluctuations over the study period, ranging from 1.4% in 2010 to 2.6% in 2014, with an overall stable trend (Table 2). A detailed year-by-year analysis revealed that the highest incidence occurred in 2014 (2.6%, n=98), followed by 2015 (2.4%, n=93) and 2013 (2.1%, n=77). In contrast, the lowest rates were observed in 2010 (1.4%, n=50) and 2016 (1.5%, n=62). Notably, no consistent upward or downward trajectory was observed across the decade, suggesting that postoperative delirium incidence remained relatively stable in this population (Fig 2).

**Fig 2.**
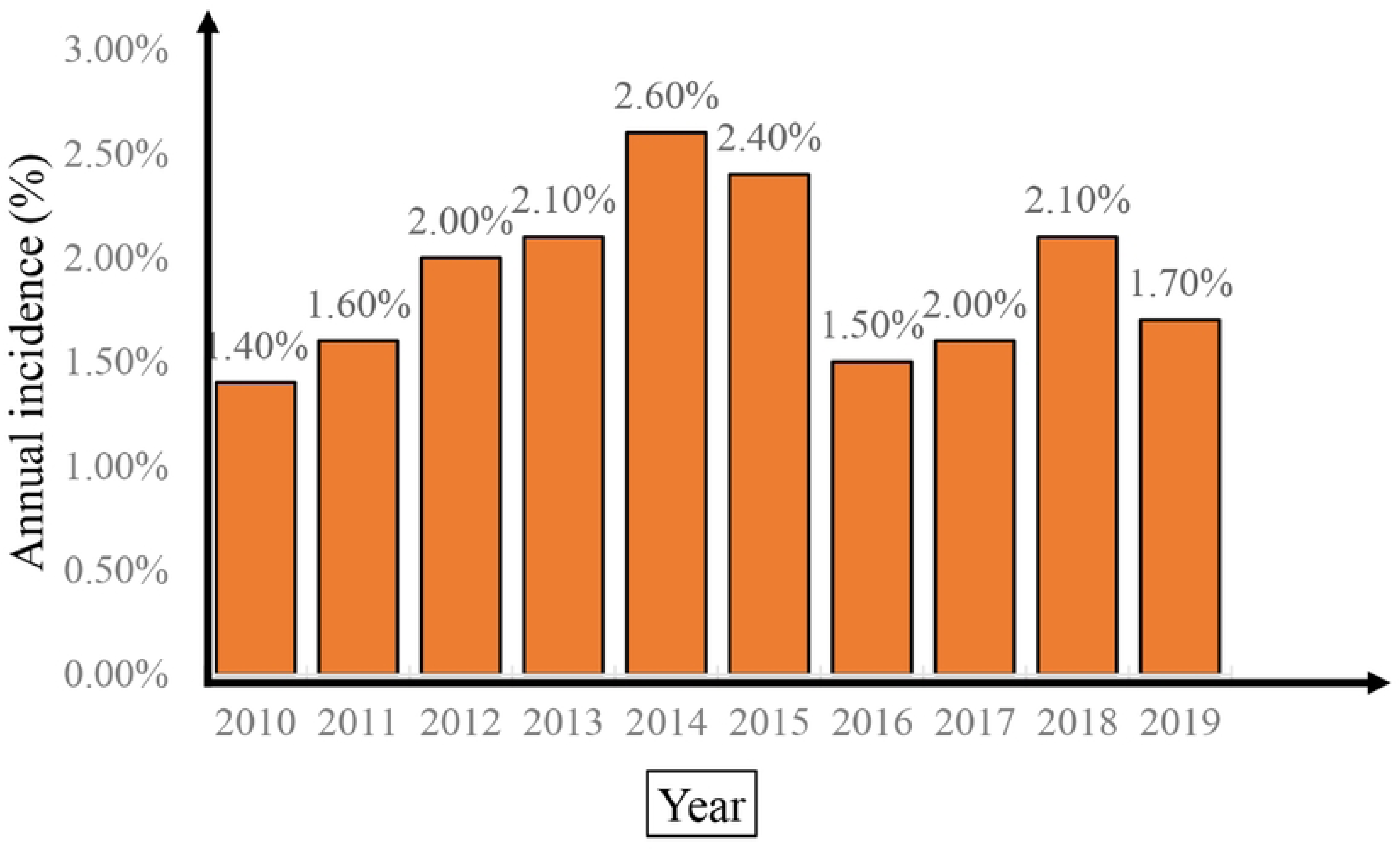
Annual Incidence of Postoperative Delirium in pediatric Patients Undergoing congenital heart disease

**Table 2.**
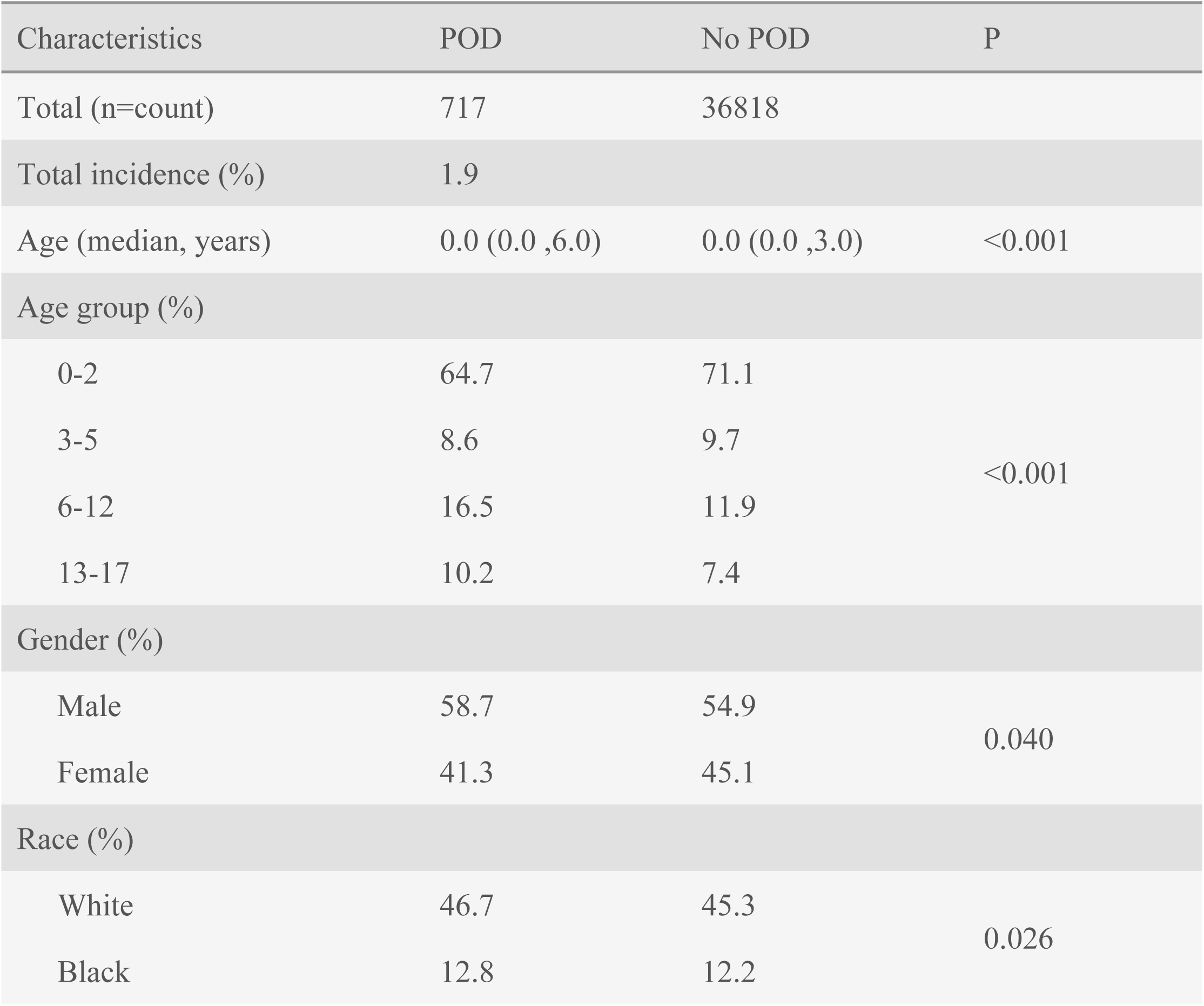
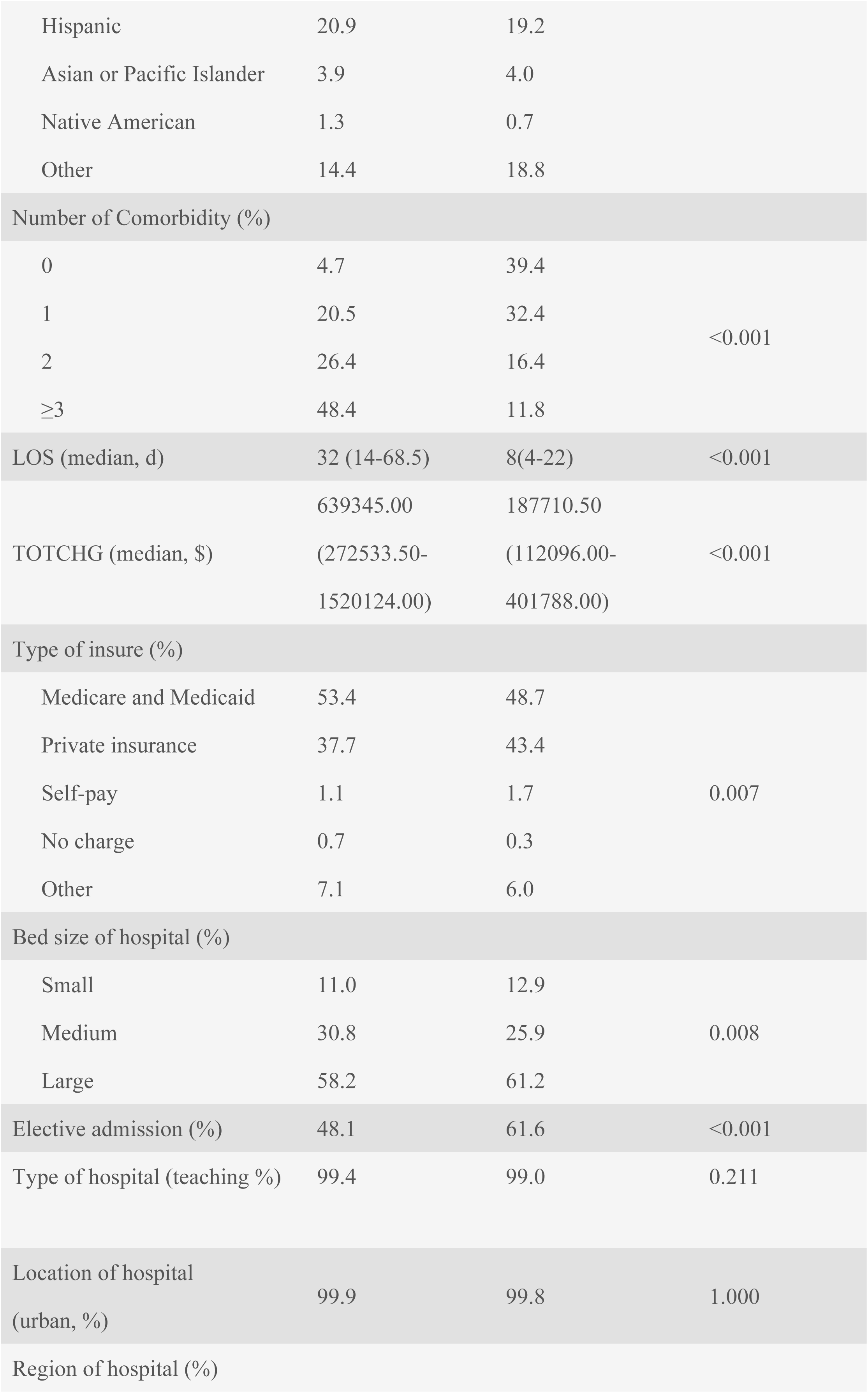
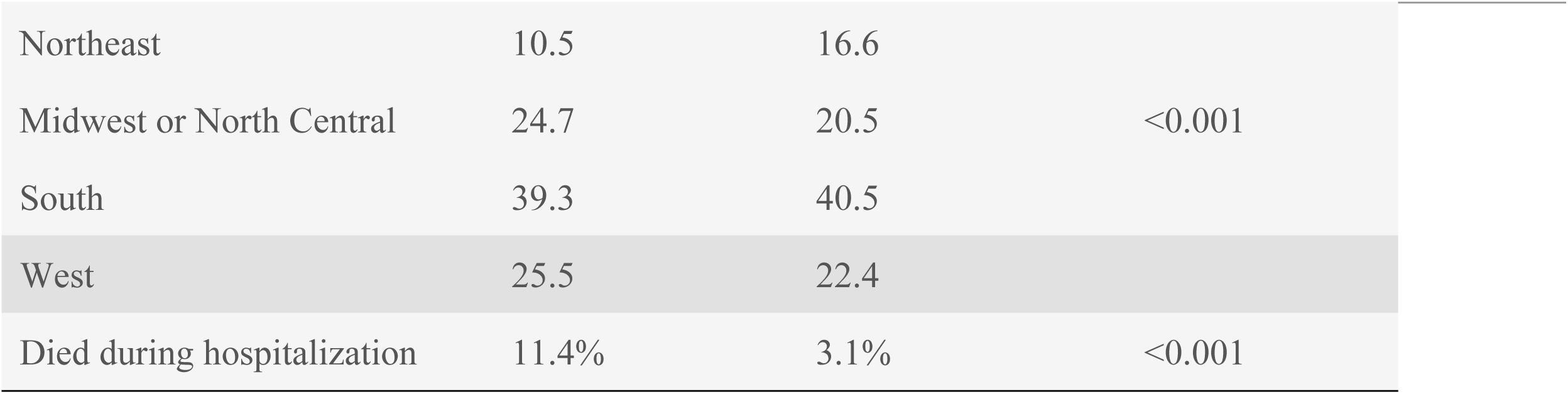
Patient characteristics and outcomes after congenital heart disease surgery (2010–2019)

### Hospital characteristics and patient demographics in two surgical groups

Significant differences were observed in age-related characteristics between POD and without POD in children undergoing congenital heart surgery. Patients with POD were significantly older (median age: 0.0 [0.0-6.0] years vs 0.0 [0.0-3.0] years, P<0.001) (Table 2). Age distribution analysis revealed markedly different patterns, with the POD group having lower proportions of infants (0-2 years: 64.7% vs 71.1%) but higher proportions of older children (6-12 years: 16.5% vs 11.9%; 13-17 years: 10.2% vs 7.4%, P<0.001) (Table 2 and Fig.3A & B).The elective admission rate was significantly lower in the POD group compared to non-POD patients (48.1% vs 61.6%, P<0.001) (Table 2). Regional variations were also significant, with higher POD proportions in the Midwest/North Central (24.7% vs 20.5%) and West (25.5% vs 22.4%) regions (P<0.001) (Table 2 and Fig.3C & D). However, there were no statistical differences in variables such as race, gender, insurance type, hospital type, region of hospital, bed size of hospital, etc. (Table 2 and Fig.4 A & B & C and D).

**Fig 3.**
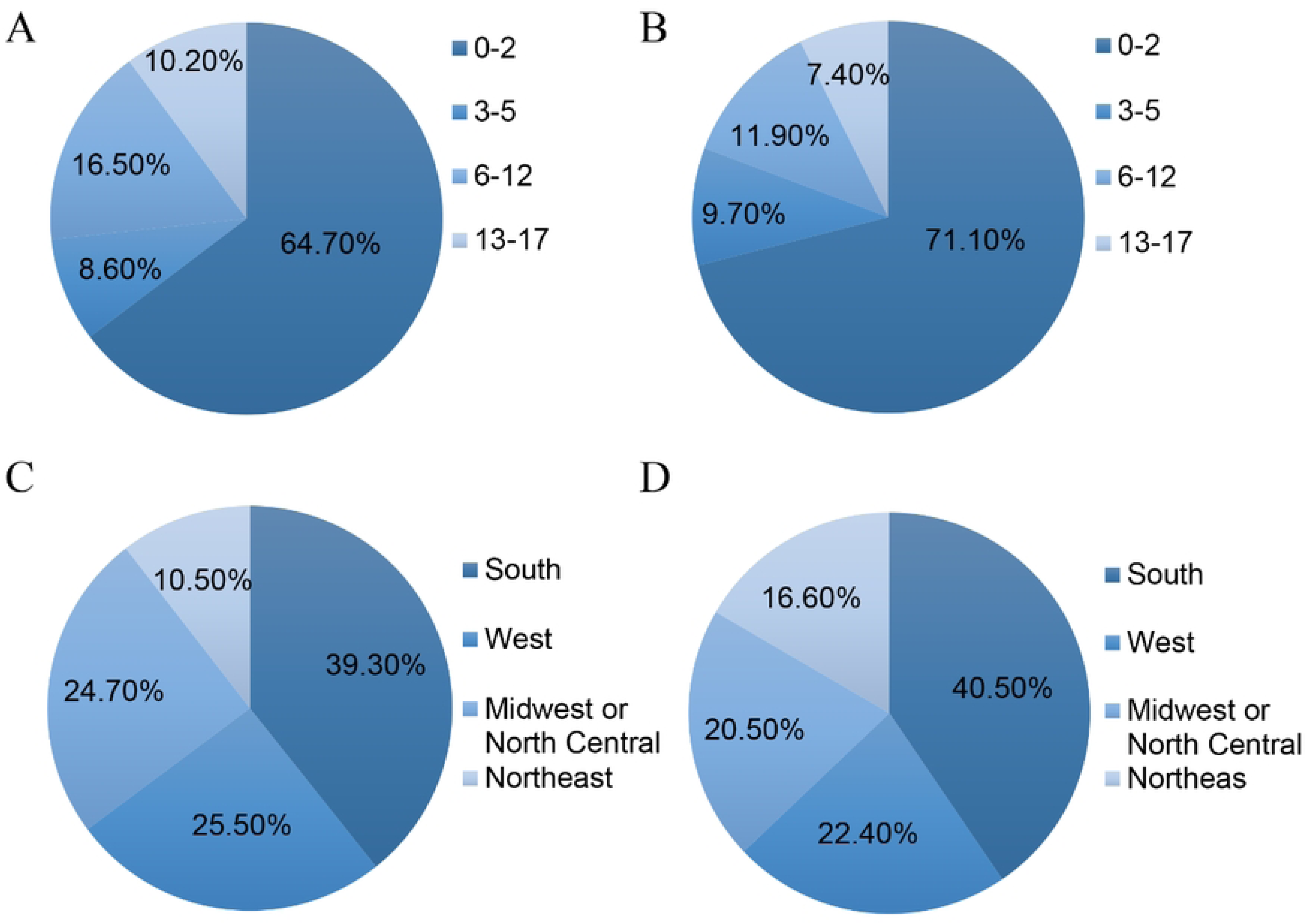
Patient demographics between the two surgical groups. A: Age distribution analysis of postoperative delirium patients. B: Analysis of age distribution of patients without postoperative delirium. C: Hospital Region of patients with postoperative delirium. D: Hospital Region of patients without postoperative delirium.

**Fig 4.**
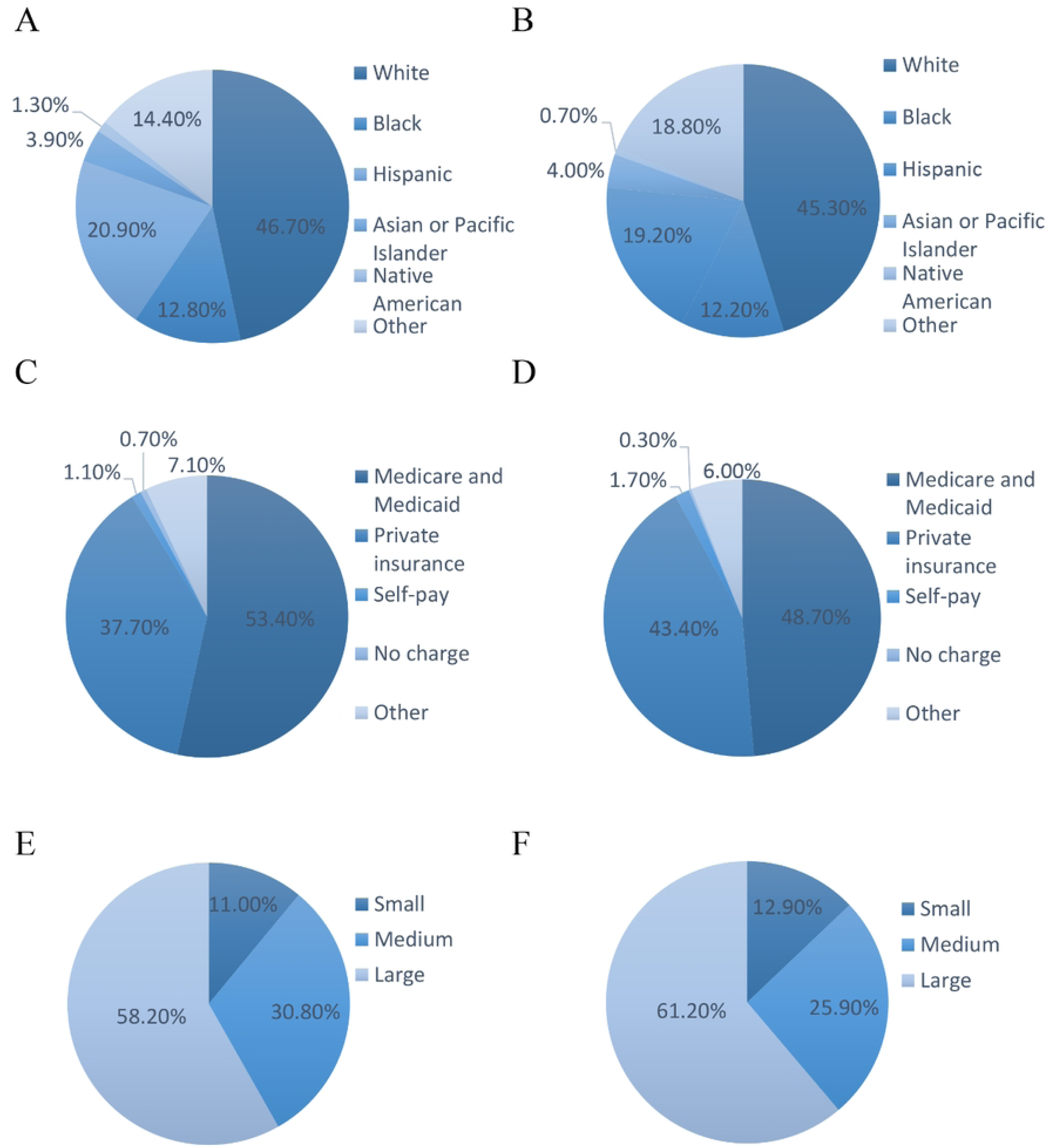
Racial distribution and incidence of Postoperative Complications Related to Postoperative Delirium. A Racial distribution analysis of delirium patients. B Racial distribution analysis of non-delirium patients. C Analysis of Insurance Types for Patients with Delirium. D Analysis of Insurance Types for Non-delirium Patients. E Analysis of the number of hospital beds for patients with delirium. F Analysis of the number of hospital beds for non-delirium patients.

### Adverse outcomes of postoperative delirium after CHD in Children

CHD children with POD demonstrated a significantly greater comorbidity burden, with 48.40% presenting ≥3 coexisting conditions compared to 11.80% in non-POD cases (P < 0.001) (Table 2). The mortality rate tripled in the delirium group (11.4% vs. 3.1%; P < 0.001) (Table 2). Hospitalization duration showed marked prolongation, with median length of stay (LOS) extending fourfold in POD patients (32 days vs. 8 days; P < 0.001) (Table2). This clinical deterioration translated into substantial economic consequences, as the total charges of hospitalization increased by $451,634.5 ($639,345 vs. $187,710.5, P < 0.001) (Table 2)

### Risk Factors of POD Following Surgery for Congenital Heart Disease in Children

Logistic regression analysis identified several significant predictors of POD in pediatric CHD surgical patients, revealing the following indicators: older age groups (6-12 years, OR=1.613; CI=1.300–2.001; P<0.001), increased comorbidity burden (1 comorbidity: OR=5.129; CI=3.529–7.456; P<0.001; 2 comorbidities: OR=12.997; CI=9.004–18.759; P<0.001;3 comorbidities: OR=31.436; CI=22.022–44.873; P<0.001) (Table 3). comorbidities including, coagulopathy (OR=2.147; CI=1.724–2.675; P<0.001), drug abuse (OR=50.18; CI=39.558–63.665; P<0.001), fluid and electrolyte disorders (OR=1.595; CI=1.334–1.908; P<0.001), other neurological disorders (OR=35.21; CI=29.115–42.598; P<0.001) and Paralysis (OR=2.182; CI=1.418-3.358; P<0.001) (Table 4).

**Table 3.** Risk factors associated with postoperative delirium following Surgery for Congenital Heart Disease in Children.

| Variable | Multivariate Logistic Regression |  |  |
| --- | --- | --- | --- |
|  | OR | 95% CI | P |
| Age group (%) |  |  |  |
| 0-2 | Ref | — | — |
| 3-5 | 1.434 | 1.082–1.899 | 0.012 |
| 6-12 | 1.613 | 1.300–2.001 | <0.001 |
| 13-17 | 1.367 | 1.054–1.774 | 0.019 |
| Female | 0.855 | 0.733-0.996 | 0.044 |
| Race |  |  |  |
| White | Ref | — | — |
| Black | 0.906 | 0.709-1.157 | 0.429 |
| Hispanic | 0.924 | 0.749-1.141 | 0.465 |
| Asian or Pacific Islander | 0.846 | 0.567-1.263 | 0.414 |
| Native American | 1.567 | 0.780-3.147 | 0.207 |
| Other | 0.843 | 0.669-1.062 | 0.148 |
| Number of Comorbidity |  |  |  |
| 0 | Ref | — | — |
| 1 | 5.129 | 3.529-7.456 | <0.001 |
| 2 | 12.997 | 9.004-18.759 | <0.001 |
| ≥3 | 31.436 | 22.022-44.873 | <0.001 |
| Type of insurance |  |  |  |
| Medicare/ Medicaid | Ref | — | — |
| Private insurance | 0.847 | 0.715-1.003 | 0.055 |
| Self-pay | 0.680 | 0.332-1.392 | 0.291 |
| No charge | 2.780 | 1.092-7.082 | 0.032 |
| Other | 0.972 | 0.714-1.325 | 0.859 |

| Bed size of hospital |  |  |  |
| --- | --- | --- | --- |
| Small | Ref | — | — |
| Medium | 1.730 | 1.314-2.278 | <0.001 |
| Large | 1.308 | 1.010-1.693 | 0.042 |
| Elective admission | 0.634 | 0.540-0.744 | <0.001 |
| Teaching hospital | 1.980 | 0.623-6.294 | 0.247 |
| Urban hospital | 0.313 | 0.031-3.176 | 0.326 |
| Region of hospital |  |  |  |
| Northeast | Ref | — | — |
| Midwest or North Central | 1.634 | 1.231-2.169 | 0.001 |
| South | 1.347 | 1.033-1.758 | 0.028 |
| West | 1.588 | 1.196-2.108 | 0.001 |

**Table 4.**
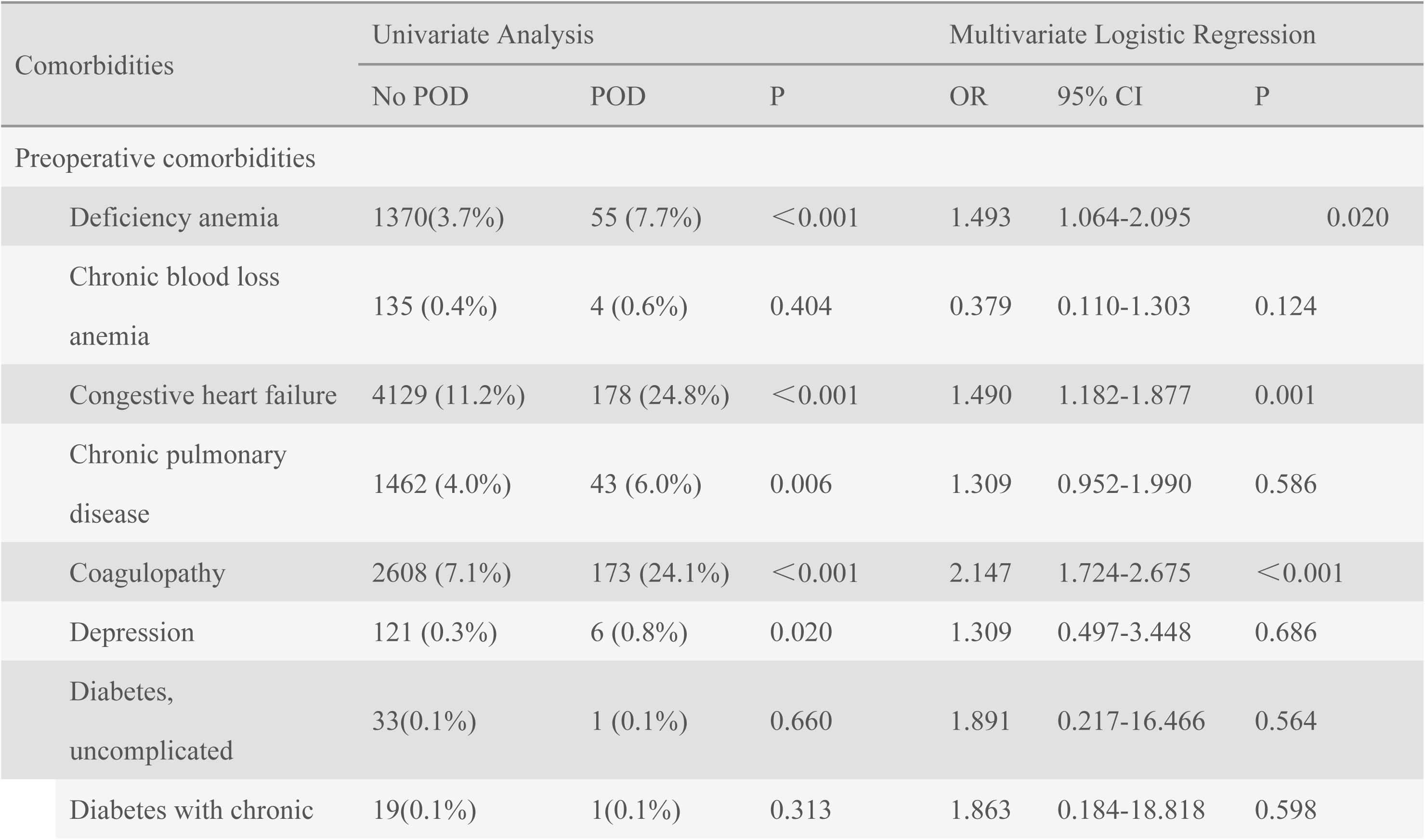
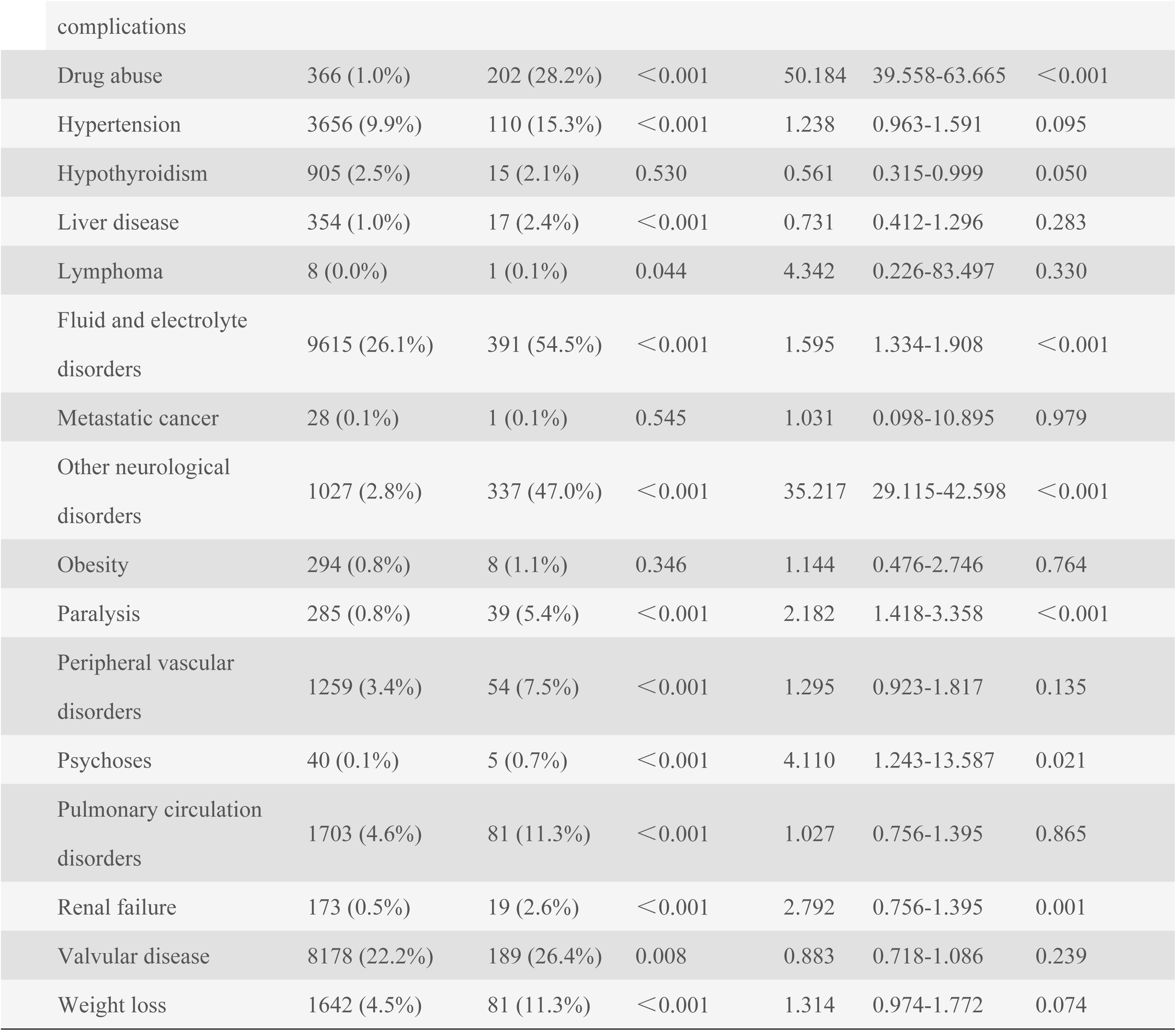
Relationship between postoperative Delirium and preoperative comorbidities.

### Risk factors associated with postoperative delirium after CHD surgery

POD occurred more frequently in pediatric CHD patients with acute myocardial infarction, acute renal failure, pneumonia, urinary tract infection, continuous trauma ventilation, arrhythmia, wound dehiscence/non-healing, postoperative shock and respiratory failure (*P* < 0.001) (Table 5). Multivariable regression analysis revealed a significant association between POD and acute renal failure (OR = 3.314; 95% CI = 2.768–3.969; P < 0.001), acute myocardial infarction (OR = 1.754; CI = 1.344–2.288; P < 0.001), pneumonia (OR = 3.431; CI = 2.763–4.262; P < 0.001), continuous trauma ventilation (OR = 1.905; CI = 1.614–2.248; P < 0.001), and respiratory failure (OR = 2.114; CI = 1.788–2.500; P < 0.001) (Table 5).

**Table 5.**
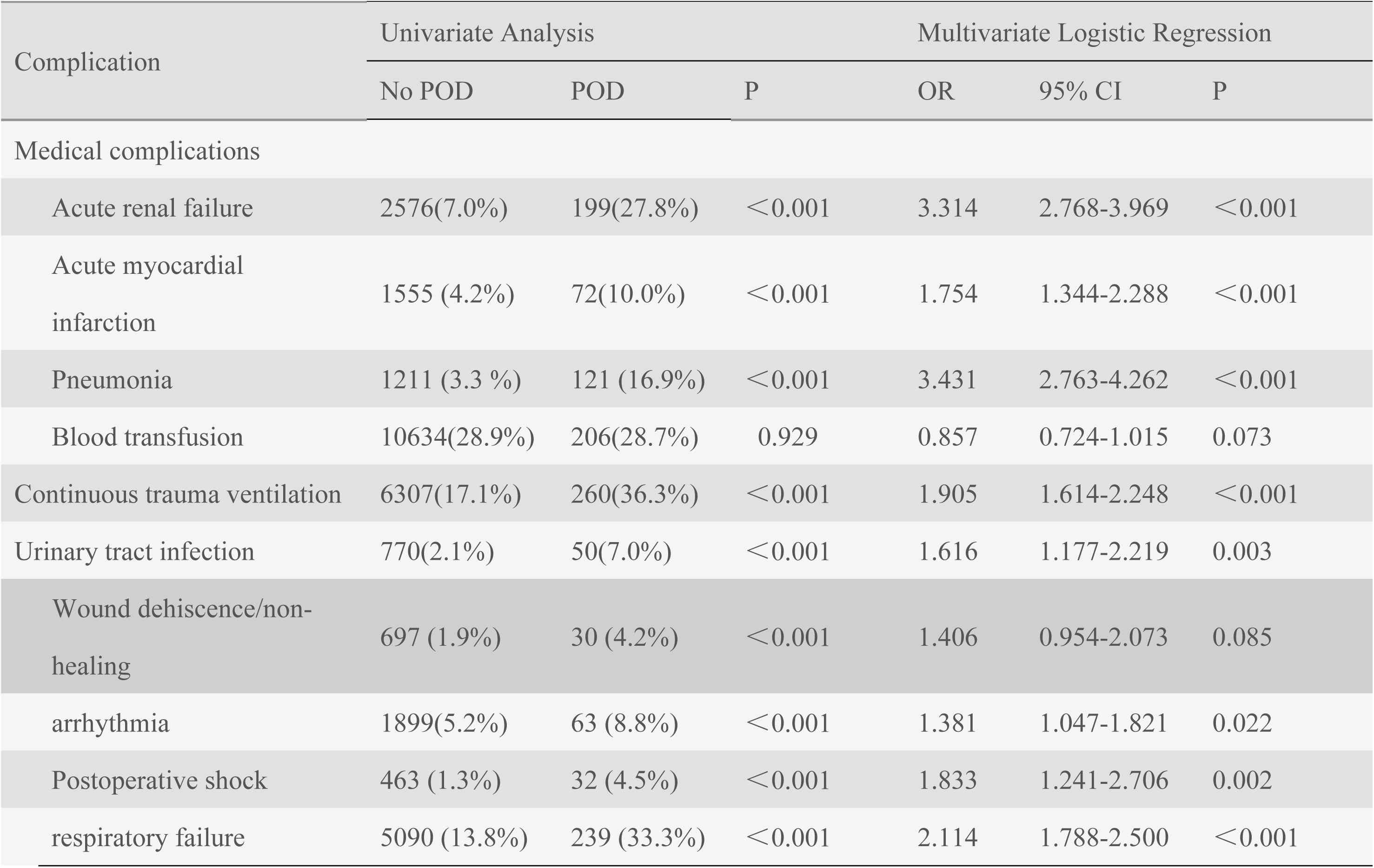
Relationship between postoperative delirium and postoperative complications.

## Discussion

POD represents a significant complication in pediatric CHD surgery that substantially impacts recovery outcomes. This large-scale health economic analysis characterizes POD epidemiology across pediatric CHD surgical populations. Our study identified an overall POD incidence of 1.9% between 2010-2019, demonstrating temporal stability. our findings contrast markedly with Mao et al.’s single-center study reporting 25.4% POD incidence following pediatric CHD surgery(16). This disparity likely reflects fundamental methodological differences between multicenter administrative data analysis and prospective clinical assessments. The ICD diagnostic criteria demonstrate more stringent parameters relative to DSM psychiatric classifications, leading to lower case identification rates(20, 21). Several inherent limitations of the NIS database contribute to potential underestimation of delirium incidence. The database exclusively captures in-hospital delirium cases and demonstrates high specificity with correspondingly low sensitivity, creating systematic underdetection(20–22). Furthermore, ICD-9-CM diagnostic codes cannot adequately document critical delirium manifestations including disorganized thinking, altered consciousness, cognitive deficits, and perceptual disturbances(23). These methodological constraints collectively suggest that our reported incidence rates represent conservative estimates of true POD prevalence in this vulnerable population.

Regarding patient demographics, our analysis identified older age as a significant risk factor for POD following CHD surgery in the pediatric population. Children aged 6 years and above constituted a substantially higher proportion of the delirium cohort compared to the non-delirium group, emerging as an independent predictor of POD development. Ours finding contradicts previous reports demonstrating that younger age was associated with higher likelihood of delirium development(7, 16, 24). We attribute this discrepancy to fundamental methodological differences: earlier single-center studies conducted predominantly at tertiary referral institutions enrolled populations skewed toward neonates with critical CHD requiring emergent intervention. In contrast, the nationwide NIS database captures a broader spectrum of school-aged pediatric patients undergoing non-emergent elective procedures across community hospitals and academic medical centers. This population-representative sampling includes substantial proportions of school-aged children undergoing elective procedures, thereby more accurately reflecting real-world epidemiological distributions.

Our analysis further revealed that preexisting comorbidities substantially increased the risk of postoperative delirium in this population. Comorbid conditions may predispose pediatric patients to heightened physiological stress and chronic inflammation, which when combined with the surgical stress-induced inflammatory response, can potentially trigger neuroinflammation and precipitate postoperative delirium(25–27). POD has been demonstrated to significantly extend length of hospital stay, escalate healthcare expenditures, and adversely impact patient survival outcomes (9, 25, 28, 29). Our investigation yielded consistent results with these previous observations. Studies have demonstrated that POD is significantly associated with prolonged hospitalization, likely attributable to the extended monitoring and specialized care required for delirious patients(9, 30). Moreover, POD increases healthcare costs through multiple mechanisms, including heightened consumption of medical resources (e.g., frequent monitoring, pharmacological therapies, and nursing interventions) and the necessity of additional treatments for delirium-associated complications such as infections or respiratory failure(28, 31).Most concerningly, POD was correlated with elevated postoperative mortality, potentially mediated by delirium-induced life-threatening complications (e.g., infections, organ dysfunction) or heightened exposure to iatrogenic risks during the prolonged hospitalization(6, 32).

Interestingly, Our regression analysis identified elective admission as a significant protective factor against POD, corroborating existing literature that demonstrates reduced delirium risk in electively admitted patients (33, 34). This protective effect may be attributed to patients’ more stable baseline health status and the opportunity for comprehensive preoperative optimization in elective cases. The likelihood of POD was lower in the Northeast and South regions but higher in the Midwest/North Central and West regions. The exact etiology remains uncertain but is probably multifactorial in nature(4). Logistic regression analysis revealed a statistically significant increase in POD rates among medium-scale facilities versus small-scale institutions. Medium hospitals may demonstrate enhanced capability for early identification of high-risk pediatric patients and implementation of preventive measures(35).

Existing evidence from pediatric cardiac surgery research indicates that implementing systematic preoperative screening, establishing validated risk stratification protocols, and applying standardized management approaches are essential components for optimizing postoperative outcomes in children with congenital heart disease(4, 9). Consequently, comprehensive evaluation of identifiable preoperative risk factors represents a fundamental prerequisite for effective prevention of postoperative delirium in this vulnerable patient population. Our logistic regression analysis produced findings that align with existing evidence from previous studies(36–40). Consistent with our hypothesis, pre-existing neurological disorders demonstrated the strongest association with POD (OR = 35.21), underscoring the vulnerability of children with preexisting neurological conditions to postoperative neuroinflammation and delirium. Similarly, drug abuse (OR = 50.18) and coagulopathy (OR = 2.147) emerged as strong independent risk factors, likely due to their systemic impact on cerebral perfusion and cerebral neuronal injury(41, 42). In addition, This study has confirmed that fluid and electrolyte disturbances (OR = 1.595) are also related to the occurrence of POD (14). Furthermore, to the authors’ knowledge, paralysis (OR = 2.182) was identified for the first time as an independent risk factor for POD. These findings align with prior adult studies linking neurological and metabolic derangements to POD, though our study highlights their amplified effects in pediatric populations.

Our retrospective analysis demonstrates significant associations between POD and specific perioperative complications following CHD surgery, including acute renal failure(OR = 3.314), acute myocardial infarction(OR =1.754), pneumonia(OR = 3.431), continuous trauma ventilation(OR = 1.905), respiratory failure(OR = 2.114). While our study identified statistically significant correlations between POD and perioperative complications in congenital heart surgery patients, the observational nature of this retrospective analysis precludes definitive causal attribution. Inherent limitations in database studies, including potential confounding variables and temporal ambiguity, prevent causal inference. These findings necessitate prospective validation through controlled longitudinal studies incorporating serial neurological assessments and stan.

While our study benefits from a large, nationally representative sample and multivariate adjustment for confounders, several limitations inherent to the NIS database must be acknowledged. First, the NIS only captures in-hospital data, excluding post-discharge complications and long-term outcomes. This may lead to underestimation of POD incidence, as delayed-onset cases are unrecorded. Second, as an administrative database, the NIS lacks granular clinical details such as surgical duration, anesthesia type (e.g., volatile vs. intravenous agents), and specific perioperative medications (e.g., opioid dosing, benzodiazepine use) — all established POD risk factors. Additionally, delirium assessment methods varied across hospitals, relying solely on ICD codes rather than standardized diagnostic tools, potentially affecting detection sensitivity. Furthermore, coding discrepancies and documentation errors, common in large databases, may introduce misclassification bias. While the NIS provides high specificity for recorded complications, its low sensitivity could result in underreporting of true POD cases. Despite these constraints, the NIS remains valuable for analyzing rare outcomes in pediatric cardiac surgery due to its extensive sample size and generalizability. Future prospective studies incorporating detailed perioperative variables and standardized delirium assessments are needed to validate these findings.

## Conclusions

POD represents a significant complication in pediatric CHD surgery, with an overall incidence of 1.9% (2010–2019). Key risk factors included older age, preoperative neurological disorders, drug abuse, and coagulopathy, while elective admission emerged as a protective factor, reflecting optimized preoperative management. Hospital characteristics revealed geographic disparities (higher Midwest/West incidence) and elevated rates at large institutions, suggesting variations in detection practices or resource allocation. POD was associated with prolonged hospitalization, increased costs, and 10-fold higher mortality, driven by complications such as acute renal failure and pneumonia. Paralysis was newly identified as a pediatric-specific risk factor, extending adult neuroinflammatory models to children.

## Data Availability

The data used in this study were obtained from the Nationwide Inpatient Sample (NIS) database, part of the Healthcare Cost and Utilization Project (HCUP) sponsored by the Agency for Healthcare Research and Quality. The NIS is a large, publicly available all-payer inpatient care database in the United States, accessible through the HCUP website at https://hcup-us.ahrq.gov/nisoverview.jsp

## Acknowledgments

We would like to thank the research participants for their unreserved help and all those who supported us, whether directly or indirectly.

## Supporting information

Supplemental Table S1. POD was diagnosed using particular ICD-9-CM and ICD-10-CM diagnostic codes.

## Notes

### Competing Interest Statement

The authors have declared no competing interest.

### Funding Statement

Yes

### Author Declarations

This study used de-identified data from the Healthcare Cost and Utilization Project (HCUP) Nationwide Inpatient Sample (NIS) database, a publicly available dataset that complies with the US Health Insurance Portability and Accountability Act (HIPAA) privacy rules. As a retrospective analysis of pre-existing anonymized data, this study was exempt from institutional review board (IRB) approval according to 45 CFR 46.104(d)(4)

## References

1. Global, regional, and national burden of congenital heart disease, 1990-2017: a systematic analysis for the Global Burden of Disease Study 2017. Lancet Child Adolesc Health. 2020;4(3):185–200.

2. Dotson A, Covas T, Halstater B, Ragsdale J. Congenital Heart Disease. Prim Care. 2024;51(1):125–42.

3. Murray MT, Krishnamurthy G, Corda R, Turcotte RF, Jia H, Bacha E, et al. Surgical site infections and bloodstream infections in infants after cardiac surgery. J Thorac Cardiovasc Surg. 2014;148(1):259–65.

4. Ghasemi Shayan R, Fatollahzadeh Dizaji M, Sajjadian F. Surgical and postoperative management of congenital heart disease: a systematic review of observational studies. Langenbecks Arch Surg. 2025;410(1):113.

5. Zmora R, Spector L, Bass J, Thomas A, Knight J, Lakshminarayan K, et al. Procedure-Specific Center Volume and Mortality After Infantile Congenital Heart Surgery. Ann Thorac Surg. 2023;116(3):525–31.

6. Patel AK, Biagas KV, Clarke EC, Gerber LM, Mauer E, Silver G, et al. Delirium in Children After Cardiac Bypass Surgery. Pediatr Crit Care Med. 2017;18(2):165–71.

7. Köditz H, Drouche A, Dennhardt N, Schmidt M, Schultz M, Schultz B. Depth of anesthesia, temperature, and postoperative delirium in children and adolescents undergoing cardiac surgery. BMC Anesthesiol. 2023;23(1):148.

8. El-Gabalawy R, Patel R, Kilborn K, Blaney C, Hoban C, Ryner L, et al. A Novel Stress-Diathesis Model to Predict Risk of Post-operative Delirium: Implications for Intra-operative Management. Front Aging Neurosci. 2017;9:274.

9. Fu M, Yuan Q, Yang Q, Song W, Yu Y, Luo Y, et al. Risk factors and incidence of postoperative delirium after cardiac surgery in children: a systematic review and meta-analysis. Ital J Pediatr. 2024;50(1):24.

10. Mao D, Fu L, Zhang W. Risk Factors and Nomogram Model of Postoperative Delirium in Children with Congenital Heart Disease: A Single-Center Prospective Study. Pediatr Cardiol. 2024;45(1):68–80.

11. Alvarez RV, Palmer C, Czaja AS, Peyton C, Silver G, Traube C, et al. Delirium is a Common and Early Finding in Patients in the Pediatric Cardiac Intensive Care Unit. J Pediatr. 2018;195:206–12.

12. Dervan LA, Di Gennaro JL, Farris RWD, Watson RS. Delirium in a Tertiary PICU: Risk Factors and Outcomes. Pediatr Crit Care Med. 2020;21(1):21–32.

13. Traube C, Mauer EA, Gerber LM, Kaur S, Joyce C, Kerson A, et al. Cost Associated With Pediatric Delirium in the ICU. Crit Care Med. 2016;44(12):e1175–e9.

14. Yang Q, Fu J, Pan X, Shi D, Li K, Sun M, et al. A retrospective analysis of the incidence of postoperative delirium and the importance of database selection for its definition. BMC Psychiatry. 2023;23(1):88.

15. Yang Q, Wang J, Huang X, Xu Y, Zhang Y. Incidence and risk factors associated with postoperative delirium following primary elective total hip arthroplasty: a retrospective nationwide inpatient sample database study. BMC Psychiatry. 2020;20(1):343.

16. Mao D, Fu L, Zhang W. Construction and validation of an early prediction model of delirium in children after congenital heart surgery. Transl Pediatr. 2022;11(6):954–64.

17. Liu K, Lin N, Jin T, Xiang Y, Li J, Lai D, et al. Association between pediatric postoperative delirium and regional cerebral oxygen saturation: a prospective observational study. BMC Psychiatry. 2024;24(1):367.

18. Zhao Y, Guo S, Wang Z, Dong Y, Wei W, Su Z. Clinical investigation into risk factors for delirium post-cardiac surgery and its implications for nursing intervention guided by behavior change theory. J Cardiothorac Surg. 2024;19(1):608.

19. Yu H, Simpao AF, Ruiz VM, Nelson O, Muhly WT, Sutherland TN, et al. Predicting pediatric emergence delirium using data-driven machine learning applied to electronic health record dataset at a quaternary care pediatric hospital. JAMIA Open. 2023;6(4):ooad106.

20. Bozic KJ, Bashyal RK, Anthony SG, Chiu V, Shulman B, Rubash HE. Is administratively coded comorbidity and complication data in total joint arthroplasty valid? Clin Orthop Relat Res. 2013;471(1):201–5.

21. Lin RY, Heacock LC, Fogel JF. Drug-induced, dementia-associated and non-dementia, non-drug delirium hospitalizations in the United States, 1998-2005: an analysis of the national inpatient sample. Drugs Aging. 2010;27(1):51–61.

22. Menendez ME, Ring D, Barnes CL. Inpatient Dislocation After Primary Total Hip Arthroplasty. J Arthroplasty. 2016;31(12):2889–93.

23. Lin RY, Heacock LC, Fogel JF. Drug-induced, dementia-associated and non-dementia, non-drug delirium hospitalizations in the United States, 1998-2005: an analysis of the national inpatient sample. Drugs Aging. 2010;27(1):51–61.

24. Lyu J, Jia Y, Yan M, Zhao Y, Liu Y-F, Li Y-L, et al. Risk factors for postoperative delirium in children with congenital heart disease: a prospective nested case-control study. Zhongguo Dang Dai Er Ke Za Zhi. 2022;24(3):232–9.

25. Mahmoudi H, Chalkias A, Moradi A, Moradian ST, Amouzegar SMR, Vahedian-Azimi A. Evaluation of postoperative delirium in cardiac surgery patients with the SDACS screening tool: a multicenter-multiphase study. Perioper Med (Lond). 2025;14(1):37.

26. Spiropoulou E, Samanidis G, Kanakis M, Nenekidis I. Risk Factors for Acute Postoperative Delirium in Cardiac Surgery Patients >65 Years Old. J Pers Med. 2022;12(9).

27. AlDaithan A, Shaheen N, Alahmari E, Smari AA, Al Ahmadi A, Almalahi A, et al. Age-specific vulnerability and high prevalence of delirium in pediatric intensive care based on a prospective cohort study. Sci Rep. 2024;14(1):31280.

28. Staveski SL, Pickler RH, Khoury PR, Ollberding NJ, Donnellan AL, Mauney JA, et al. Prevalence of ICU Delirium in Postoperative Pediatric Cardiac Surgery Patients. Pediatr Crit Care Med. 2021;22(1):68–78.

29. Ivkin A, Grigoriev E, Mikhailova A. Impact of Intraoperative Blood Transfusion on Cerebral Injury in Pediatric Patients Undergoing Congenital Septal Heart Defect Surgery. J Clin Med. 2024;13(20).

30. Matsuishi Y, Hoshino H, Enomoto Y, Shimojo N, Matsubara M, Kato H, et al. Pediatric delirium is associated with increased brain injury marker levels in cardiac surgery patients. Sci Rep. 2022;12(1):18681.

31. Flores AER, Oura KHU, Rocha PK, Belela-Anacleto ASC, Kusahara DM. Incidence and Factors Associated with Delirium in Children in a Single Pediatric Intensive Care Unit in Brazil. J Pediatr Nurs. 2021;61:e29–e34.

32. Koster S, Hensens AG, Schuurmans MJ, van der Palen J. Consequences of delirium after cardiac operations. Ann Thorac Surg. 2012;93(3):705–11.

33. Janssen TL, Alberts AR, Hooft L, Mattace-Raso F, Mosk CA, van der Laan L. Prevention of postoperative delirium in elderly patients planned for elective surgery: systematic review and meta-analysis. Clin Interv Aging. 2019;14:1095–117.

34. Jin Z, Hu J, Ma D. Postoperative delirium: perioperative assessment, risk reduction, and management. Br J Anaesth. 2020;125(4):492–504.

35. Mirzaaghayan M, Memarian S, Abdshah A, Ghavami M, Roozbahani G, Naseri F, et al. Delirium among pediatric patients admitted to open-heart surgery intensive care unit: A cross-sectional study investigating a common challenge and concern, and its inciting factors. Curr J Neurol. 2024;23(3):152–8.

36. Wang L-H, Xu D-J, Wei X-J, Chang H-T, Xu G-H. Electrolyte disorders and aging: risk factors for delirium in patients undergoing orthopedic surgeries. BMC Psychiatry. 2016;16(1):418.

37. Meyer M, Schwarz T, Renkawitz T, Maderbacher G, Grifka J, Weber M. Hospital Frailty Risk Score predicts adverse events in revision total hip and knee arthroplasty. Int Orthop. 2021;45(11):2765–72.

38. Peng Z, Wu J, Wang Z, Xie H, Wang J, Zhang P, et al. Incidence and related risk factors for postoperative delirium following revision total knee arthroplasty: a retrospective nationwide inpatient sample database study. BMC Musculoskelet Disord. 2024;25(1):633.

39. Liu R, Liu N, Suo S, Yang Q, Deng Z, Fu W, et al. Incidence and risk factors of postoperative delirium following hepatic resection: a retrospective national inpatient sample database study. BMC Surg. 2024;24(1):151.

40. Zheng Y, Wang J, Liu Z, Wang J, Yang Q, Ren H, et al. Incidence and Risk Factors of Postoperative Delirium in Lumbar Spinal Fusion Patients: A National Database Analysis. World Neurosurg. 2025;193:593–604.

41. Terrando N, Akassoglou K. Breaking barriers in postoperative delirium. Br J Anaesth. 2022;129(2):147–50.

42. Wu X, Zhang N, Zhou B, Liu S, Wang F, Wang J, et al. Alcohol consumption may be associated with postoperative delirium in the elderly: the PNDABLE study. BMC Anesthesiol. 2023;23(1):222.

